# Variant abundance estimation for SARS-CoV-2 in wastewater using RNA-Seq quantification

**DOI:** 10.1101/2021.08.31.21262938

**Authors:** Jasmijn A. Baaijens, Alessandro Zulli, Isabel M. Ott, Mary E. Petrone, Tara Alpert, Joseph R. Fauver, Chaney C. Kalinich, Chantal B.F. Vogels, Mallery I. Breban, Claire Duvallet, Kyle McElroy, Newsha Ghaeli, Maxim Imakaev, Malaika Mckenzie-Bennett, Keith Robison, Alex Plocik, Rebecca Schilling, Martha Pierson, Rebecca Littlefield, Michelle Spencer, Birgitte B. Simen, Yale SARS-CoV-2 Genomic Surveillance Initiative, William P. Hanage, Nathan D. Grubaugh, Jordan Peccia, Michael Baym

**Author notes:** Denotes equal contributions (Jasmijn A. Baaijens, Alessandro Zulli, Isabel M. Ott). Denotes co-senior authorship (Nathan D. Grubaugh, Jordan Peccia, Michael Baym).

## Abstract

Effectively monitoring the spread of SARS-CoV-2 variants is essential to efforts to counter the ongoing pandemic. Wastewater monitoring of SARS-CoV-2 RNA has proven an effective and efficient technique to approximate COVID-19 case rates in the population. Predicting variant abundances from wastewater, however, is technically challenging. Here we show that by sequencing SARS-CoV-2 RNA in wastewater and applying computational techniques initially used for RNA-Seq quantification, we can estimate the abundance of variants in wastewater samples. We show by sequencing samples from wastewater and clinical isolates in Connecticut U.S.A. between January and April 2021 that the temporal dynamics of variant strains broadly correspond. We further show that this technique can be used with other wastewater sequencing techniques by expanding to samples taken across the United States in a similar timeframe. We find high variability in signal among individual samples, and limited ability to detect the presence of variants with clinical frequencies <10%; nevertheless, the overall trends match what we observed from sequencing clinical samples. Thus, while clinical sequencing remains a more sensitive technique for population surveillance, wastewater sequencing can be used to monitor trends in variant prevalence in situations where clinical sequencing is unavailable or impractical.

## Introduction

As the SARS-CoV-2 pandemic continues, the virus is evolving in real time, challenging existing control measures. Increased infectivity, and potentially increased morbidity and immune evasion, have been observed in emerging *variant* lineages^1,2^. These variants are characterized by combinations of mutations compared to the original strain, some of which are likely to be selective adaptations of the SARS-CoV-2 virus^1^. To adapt our approach to the pandemic with the virus, we need first to be able to observe which variants are present where, and critically how the rates of variants are changing in the population.

Genomic surveillance of SARS-CoV-2 enables early detection and clinical investigation of emerging variants. In late 2020, the Centers for Disease Control and Prevention (CDC) designated specific viral lineages as *variants of interest* or *variants of concern* based on potential changes in detectability, transmissibility, disease severity, therapeutic efficacy, and/or ability to evade control by natural or vaccine-induced immune responses^3^. Initially, this designation included lineage B.1.1.7 (corresponding to WHO designation *Alpha*), B.1.351 (*Beta*), and P.1 (*Gamma*). In early 2021, CDC added two new variants to this list: B.1.427 and B.1.429 (*Epsilon*), both of which were first identified in California^4,5^. B.1.617.2 and related sublineages (*Delta*) now pose a threat but were not observed at substantial rates in the United States until May 2021. Each variant is identified by a set of potentially overlapping amino acid mutations, which can be identified by genome sequencing. This is typically done by sequencing remnant clinical samples used for diagnostics (e.g., nasal swabs), but as infected patients excrete high levels of SARS-CoV-2 RNA, variant prevalences are potentially detectable from domestic wastewater.

Measuring the concentration of SARS-CoV-2 in domestic wastewater can be an efficient method for indicating infection dynamics in a population^6^. SARS-CoV-2 and fragments of its RNA genome are excreted by infected individuals through feces or urine^7^ and collected in domestic wastewater. Viral RNA in wastewater can then be extracted and quantified via quantitative RT-qPCR. This approach has been used to measure SARS-CoV-2 abundance over time, across different regions^8^ and wastewater RNA concentrations are correlated with COVID-19 case rates^9,10^. The genomes of SARS-CoV-2 in wastewater can also be sequenced, which can then be used to identify mutations present in an entire community with respect to the reference genome^11,12^. Genome analysis from wastewater sequencing is particularly challenging because of the low concentrations and poor, fragmented quality of RNA, and the presence of PCR inhibiting compounds which can interfere with library preparation in wastewater. This typically yields poor quality sequencing data where the sequencing depth is highly variable across the SARS-CoV-2 genome and overall genome coverage is often incomplete. Despite these challenges, recent work has shown that it is feasible to observe mutations in wastewater sequencing data^11,12^, and suggests the possibility of monitoring the abundance of specific lineages. Throughout the world, SARS-CoV-2 wastewater surveillance has been conducted for wastewater collection systems that serve populations ranging from 10,000 to greater than 100,000 people^13^. A method for the quantitative measurement of variants in wastewater would provide a cost- and resource-efficient approach to population genome surveillance.

Here we introduce a technique to monitor for SARS-CoV-2 variants in a population by sequencing directly from wastewater and predicting abundances via a computational approach previously used for RNA-seq transcript quantification. We demonstrate the efficacy of this approach on wastewater data collected from Connecticut between January and April 2021, during the third wave of the SARS-CoV-2 pandemic, and compare these predictions to clinically observed variant frequencies from the same geographic area and time period. We then show the generality of the approach by expanding our analysis to samples collected across the United States from late December 2020 to January 2021.

## Results

### Prediction of variant abundance is computationally analogous to RNA transcript abundance estimation

SARS-CoV-2 RNA fragments in wastewater originate from different infections, with potentially different viral lineages, pooled together into a single sample. After successfully extracting RNA from wastewater and sequencing SARS-CoV-2 genome fragments, the computational challenge is to assign reads to lineages and estimate relative abundance per lineage. This is analogous to RNA transcript quantification from RNA-Seq data, where the sequencing data consists of reads originating from different transcripts of a given gene, and the objective is to quantify the relative abundance per transcript (**Fig 1a**).

**Figure 1.**
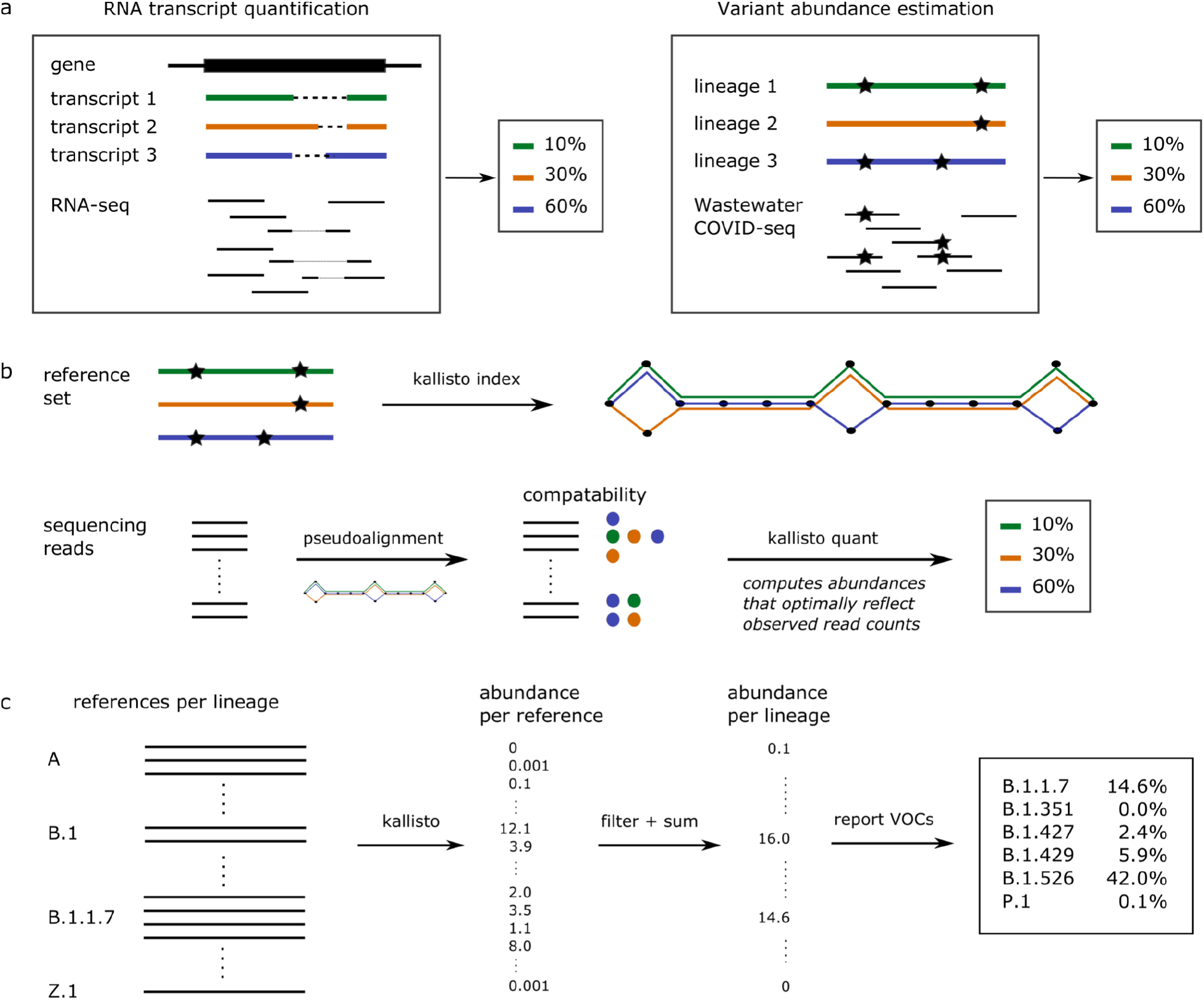
Computational approach to variant of concern (variant) abundance estimation. **a)** Computational similarity between RNA transcript quantification and variant abundance estimation. **b)** Key aspects of the kallisto algorithm in the context of variant abundance estimation. **c)** Our workflow uses multiple reference sequence per lineage to capture within-lineage variation. Applying kallisto (as in part b) results in abundance estimates per reference sequence. These abundances are filtered using a minimal abundance cutoff and subsequently summed per lineage to obtain abundance estimates per lineage. Finally, variant abundances are reported.

While pipelines for viral variant detection in clinical samples rely on individual mutation frequencies^14^ using popular tools like V-pipe^15^ or iVar^16^, RNA transcript quantification algorithms make use of the genome sequences without identifying specific mutations. This is a major advantage because identifying individual mutation frequencies from wastewater sequencing data is highly error-prone---any errors would be propagated into variant abundance estimates. Moreover, the fact that RNA transcript quantification tools have already been in use for several years in the RNA-Seq community has resulted in well-developed, user-friendly software that can be applied almost immediately to the variant abundance quantification problem.

Here, we predict variant abundance by applying kallisto^17^. This algorithm takes as input a set of reference sequences to be quantified: for RNA-Seq these would be the different transcripts, but for wastewater sequencing data we provide it with a collection of SARS-CoV-2 genomes representative of the population. kallisto constructs an index from the reference sequences and subsequently matches sequencing reads to references, which allows it to estimate the abundance of each reference transcript provided (**Fig 1b**). SARS-CoV-2 lineages are characterized by a combination of mutations, but additional variation is observed within lineages (**Fig S1**). We provide kallisto with a reference set consisting of multiple genomic sequences per lineage, capturing the mutations specific to this lineage as well as within-lineage variation (**Fig 1c**). Our constructed reference set includes 1-17 genome sequences per lineage, with a total number of nearly 1500 sequences for the 881 unique SARS-CoV-2 lineages present in the GISAID database at the time of download (9 March 2021). Including multiple sequences per lineage reduces biases related to within-lineage variation and potentially identifies any additional genomic signatures frequently seen in a given lineage. Finally, we filter out any predictions below a given percent abundance threshold to reduce noise and sum all predictions per lineage, which gives predicted abundances per lineage (**Fig 1c**). While in this study we used kallisto, we expect similar results with comparable tools such as salmon^18^.

### Kallisto predictions of variant abundance are accurate on simulated data

To evaluate the accuracy of the predictions obtained through our pipeline, we created a collection of benchmarking datasets that resemble real wastewater samples. For each variant (B.1.1.7, B.1.351, B.1.427, B.1.429, P.1) we created a series of 33 benchmarks by simulating sequencing reads from a variant genome, as well as a collection of background (non-variant of concern/interest) sequences, such that the variant abundance ranges from 0.05% to 100%. Analogously, we created a second series of benchmarks, simulating reads only from the Spike gene of each SARS-CoV-2 genome. We refer to the first set of benchmarks as “whole genome” and to the second set of benchmarks as “Spike-only”. Finally, we performed these benchmarking experiments at different sequencing depths: 100x and 1000x coverage for the whole genome benchmarks, and 100x, 1000x, and 10,000x coverage for the Spike-only benchmarks (**Table 1 and Fig 2**).

**Table 1.** Performance statistics per dataset. Results separated by a forward slash correspond to an abundance threshold of 0.1% and 1%, respectively. FPR = false positive rate; FNR = false negative rate; relative estimation error reflects the average relative frequency estimation error across all true positives.

| Benchmark | FPR | FNR | Precision | Recall | Relative estimation error (%) |
| --- | --- | --- | --- | --- | --- |
| Whole genome 100x | 0.191 / 0.0 | 0.057 / 0.032 | 0.423 / 1.0 | 0.943 / 0.968 | 29.4 / 19.4 |
| Whole genome 1,000x | 0.163 / 0 | 0.007 / 0.042 | 0.470 / 1.0 | 0.993 / 0.958 | 27.1 / 18.5 |
| Spike-only 100x | 0.121 / 0.003 | 0.107 / 0.074 | 0.508 / 0.978 | 0.893 / 0.926 | 26.3 / 15.8 |
| Spike-only 1,000x | 0.041 / 0.003 | 0.043 / 0.042 | 0.753 / 0.978 | 0.957 / 0.958 | 17.3 / 14.0 |
| Spike-only 10,000x | 0.010 / 0 | 0.014 / 0.042 | 0.926 / 1.0 | 0.986 / 0.958 | 15.3 / 13.0 |

**Figure 2.**
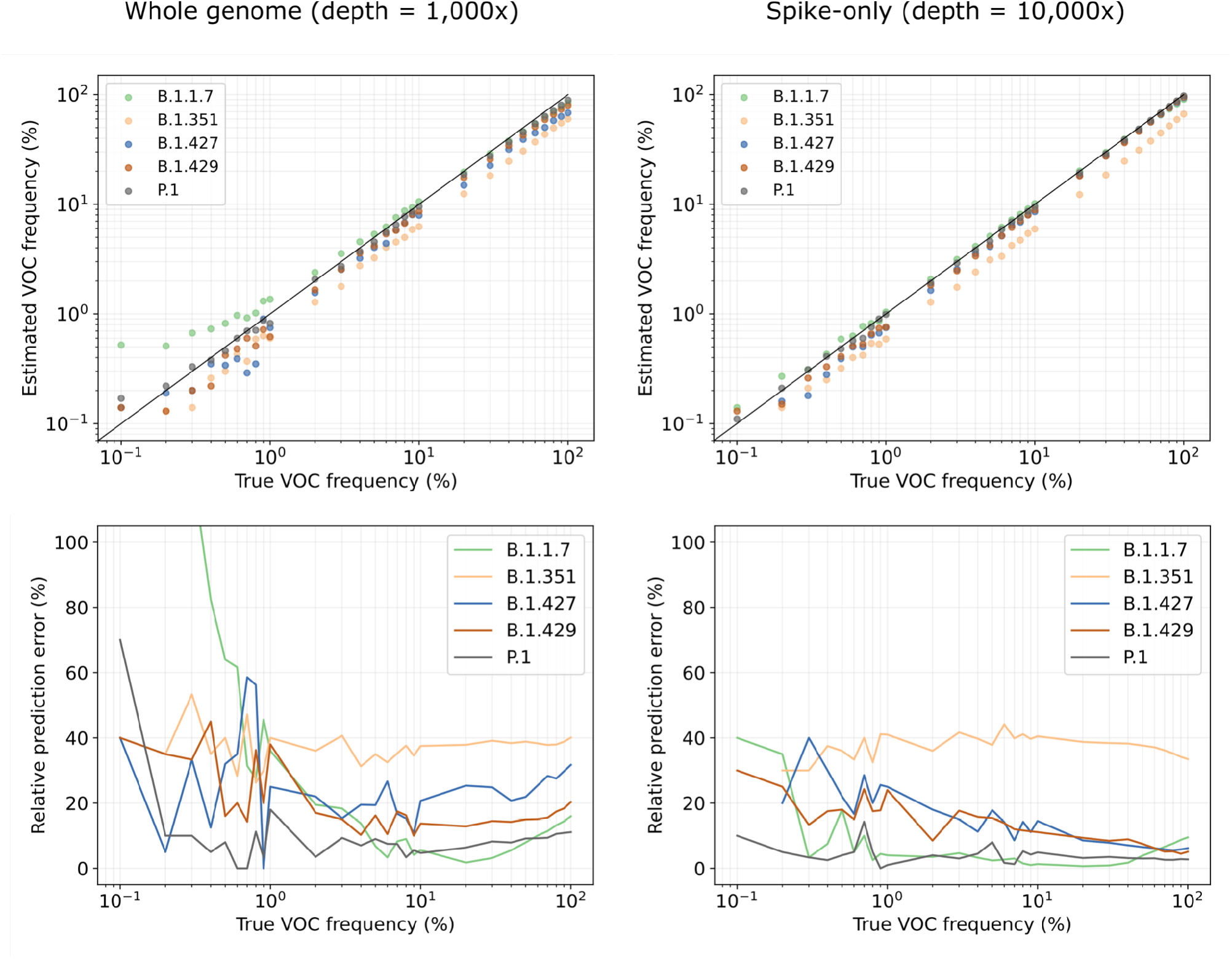
Estimated variant abundances and relative prediction errors. Relative prediction errors are defined as the absolute difference between true and estimated frequency, relative to the true frequency.

Predicting variant abundance can be difficult when a variant is present at very low frequency, because of the high degree of similarity between lineages. On our simulated datasets, where we know the true frequency of each variant, we observe a background noise of 0.01--0.09% (**Fig S2**), meaning that some sequences are falsely predicted to be present at 0.01--0.09% abundance. These false positives are likely due to shared mutations or conserved sequences between lineages. The level of background noise tends to be higher for whole genome benchmarks than for Spike-only benchmarks, because the majority of defining variant mutations are in the Spike gene (**Figs S1 and 2**). In both cases, we apply a threshold of 0.1% abundance to include a sequence in the lineage abundance computation and we only report the presence of a variant exceeding this threshold to avoid false positives. For this reason, we only report results for benchmarks with a true variant abundance of at least 0.1%. Note that this threshold applies to the overall sequence abundance and not to individual mutations, since the stochasticity of wastewater sequencing causes the abundance of mutations within a variant to vary significantly.

**Figure 2** (top) shows the predicted versus true frequencies per variant for two of the benchmarks; additional results are shown in **Figure S3**. In general, variant frequencies tend to be underestimated, in particular on whole genome data. This is another consequence of shared polymorphisms between relatively closely related lineages: a fraction of the reads is assigned to other, locally identical genomes, leading to an underestimated variant frequency. The more divergent a variant is in comparison with other lineages and the more unique polymorphisms are associated with it, the lower the number of false positives and the smaller the underestimation of variant abundance. This explains why our predictions are most accurate for P.1, the most divergent lineage among the variants considered (**Fig S1**). **Figure 2** (bottom) shows the relative frequency estimation error, defined as the absolute difference between true and estimated frequency, relative to the true frequency. We observe that relative frequency estimation errors are highest at low frequencies, where a small deviation in the absolute sense makes a large relative difference but are relatively stable at true variant frequencies of at least 1%.

Besides underestimation, we also notice overestimation, particularly at low variant frequencies. This is due to shared sequence between the variant and the background lineages: in datasets where the variant is present at a low frequency, the background lineages must be present at relatively high frequencies. Any shared sequence between a background lineage and the variant will then lead to more reads being assigned to the variant, hence overestimation. This effect only applies when background lineages with shared sequence are more abundant than the variant. In **Figure 2**, we can clearly see how P.1 and B.1.1.7 abundances are overestimated at low frequencies, while being near-perfect at higher frequencies (>1%); for other variants, we see that this effect compensates some of the underestimation, resulting in better estimates at low variant frequencies.

Two variants which are particularly difficult to predict individually are B.1.427 and B.1.429; these lineages are highly similar and have the same characterizing mutations in the Spike gene^19^. However, because we include multiple sequences per lineage in our reference set, we capture within-lineage variation that allows us to distinguish between these two lineages using Spike-only data. This highlights the power of our approach using a complete reference set; it would not be possible to distinguish between B.1.427 and B.1.429 with an approach based on mutation frequencies alone.

We observe that variant frequencies are consistently underestimated using our approach, except for slight overestimation of unusually divergent lineages (P.1, B.1.1.7) at low frequencies. Consistent variant bias as observed in our experiments is unlikely to be an issue in differential analysis, but in single-point evaluations it would be necessary to design a method to correct for these variant-specific biases. However, our benchmarks are generated using a single variant sequence, while real data consists of a mixture of different sequences for the same variant. This may have resulted in stronger underestimation on our benchmarks than would be seen on real data. To complement the benchmarking experiments presented here, it would be interesting to evaluate predictions on real data more thoroughly, e.g., by comparing to qPCR-based variant abundance estimates per variant. More extensive benchmarking experiments will make it possible to learn the variant-specific biases more accurately and adjust predictions accordingly.

To evaluate false positive and false negative predictions, we computed for each experiment the overall false positive rate (FPR), false negative rate (FNR), precision, and recall (**Table 1**). We calculated these statistics for minimal variant abundance thresholds of 0.1% and 1%. Increasing the minimal abundance thresholds reduces the false positive rate but increases the false negative rate. We generally observe more false negatives for datasets of lower coverage, because low-frequency variants become harder to detect. In terms of precision and recall, we note that, unsurprisingly, increasing the number of reads (either by amplifying a larger region, or by increased sequencing depth) leads to better results. If a variant is uniquely defined by mutations on Spike, then sequencing depth is preferred over breadth, but if a variant is (nearly) identical to other lineages on Spike, e.g., B.1.427/B.1.429, then whole genome sequencing is preferable.

While for this study we primarily used kallisto^17^, we also evaluated performance for the software package salmon^18^, which takes a slightly different algorithmic approach to the same problem (**Fig S4**) and found predictions were highly similar to those obtained with kallisto, the main difference being that salmon is slightly more conservative: it achieves higher precision (fewer false positives), at the expense of lower recall (more false negatives). Although salmon tends to miss variants at very low frequencies, one potential advantage is that this method may also be applied to long reads (in alignment-based mode), while kallisto usage is limited to short reads.

### Observed PCR values of wastewater correspond to genome coverage

We obtained primary sewage sludge samples from the wastewater treatment plant serving New Haven, CT, USA, every 2 days between January 1, 2021 and April 27, 2021 (59 samples). The observed SARS-CoV-2 RNA levels in these samples follow the same trend as COVID-19 case rates in the same geographic region (**Fig 3a**, similarly found in ^10^). PCR cycle threshold (Ct) values of SARS-CoV-2 from undiluted sludge samples ranged from 30.2 to 35.3 (7.1×10^3 - 1.6×10^5 virus copies / mL), indicating that there was enough viral RNA to apply genomic sequencing. We generated 400nt tiled amplicons encompassing the SARS-CoV-2 genome using the Illumina COVIDSeq Test (RUO), modified to use NEBNext ARTIC V3 SARS-Cov-2 Primer Mixes 1 and 2 to improve genome coverage at low RNA concentrations^20,21^. The resulting sequencing data varied widely in terms of number of reads and genome coverage (**Fig S5**), with low Ct values (high SARS-CoV-2 RNA concentrations) generally leading to higher genome coverage (**Fig 3b**). For these datasets, Ct values < 31 yields at least 60% genome coverage; samples with a Ct value < 34 and at least 0.5M reads aligned reach a genome coverage of at least 40%.

**Figure 3.**
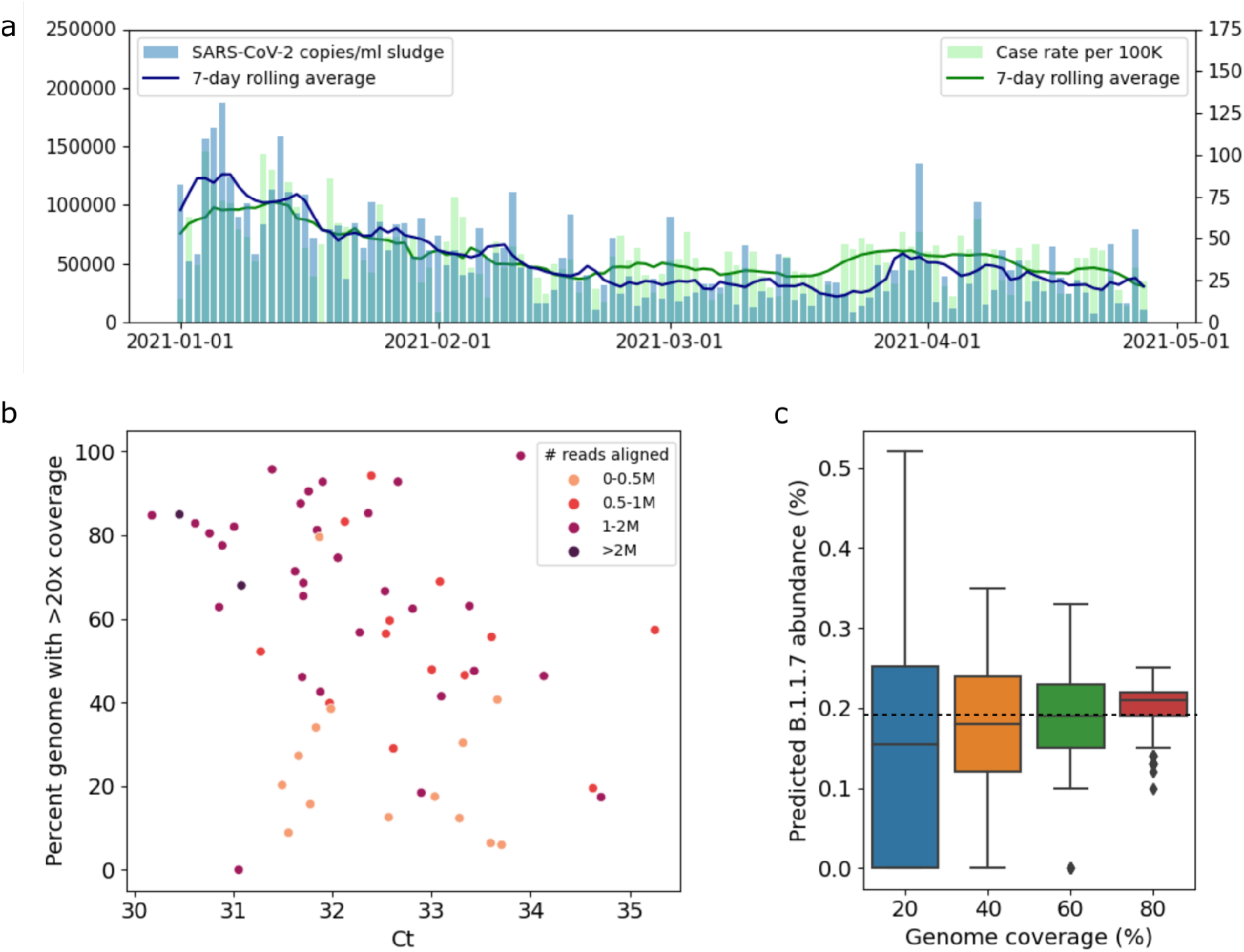
**a**) RNA levels in wastewater (copies/ml sludge, displayed on left vertical axis) follow the same trend as COVID-19 case rates (cases per 100K people, displayed on right vertical axis). **b**) Percent genome with >20x coverage versus sludge Ct values. **c**) Impact of genome coverage on predicted B.1.1.7 abundance for random subsamples of a sludge sample with full genome coverage. The horizontal dotted line indicated the predicted B.1.1.7 abundance for the full sample (99% genome coverage).

To evaluate the impact of genome coverage on variant abundance predictions, we subsampled a dataset with maximal coverage (99% of the SARS-CoV-2 genome with >20x coverage, 1.9M paired-end reads) to obtain datasets with reduced genome coverage by randomly selecting 20%, 40%, 60%, and 80% of amplicons, respectively, each of which we repeated 100 times. **Figure 3c** shows the resulting abundance predictions for B.1.1.7 per coverage value. We observe that the median predicted abundance is close to the predicted abundance at full coverage (dashed line in **Fig 3c**) for all coverage values; however, variance is much larger in datasets with low coverage compared to datasets with high coverage, consistent with statistical predictions and prior work^16^. This indicates that datasets with low genome coverage can still result in accurate abundances, but the predictions are less reliable.

### Wastewater abundances of B.1.1.7 and B.1.526 in Connecticut broadly correspond to clinically observed frequencies

We applied our abundance prediction pipeline to the series of wastewater sequencing datasets described above. **Figure 4** shows the resulting predictions for lineages B.1.1.7 and B.1.526. There is a clear trend in B.1.1.7 abundance emerging in early February 2021, increasing in abundance through mid April 2021, while the abundance of other variants is relatively stable over time (see also **Fig S6**).

**Figure 4.**
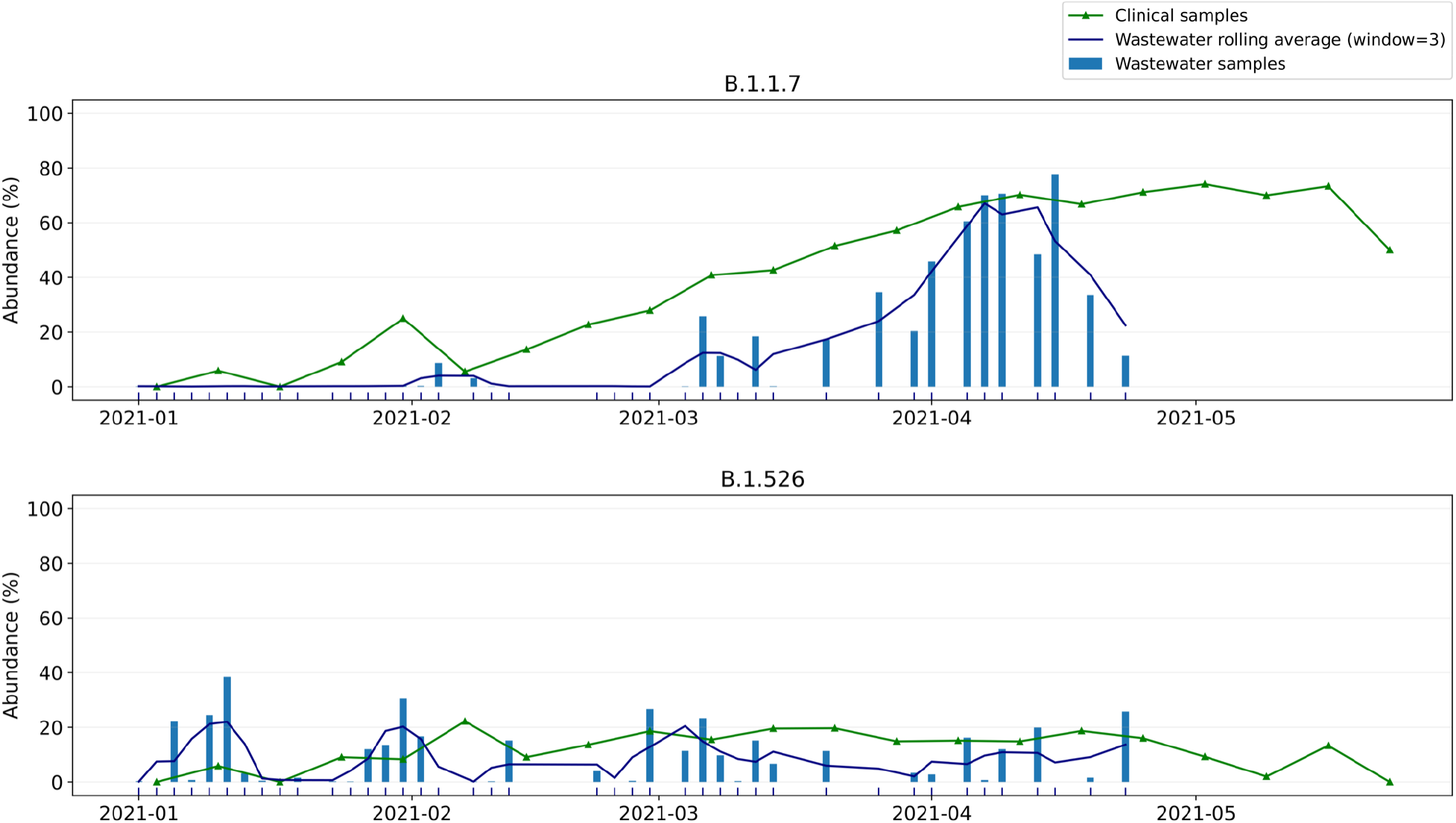
Wastewater versus clinical abundance estimates for B.1.1.7 and B.1.526 in New Haven from early January 2021 to late April 2021. Dates of clinical sampling correspond to the date of specimen collection.

We then compared our wastewater abundance predictions to variant frequency estimates from data generated by sequencing remnant clinical diagnostic samples (mostly nasal swabs) in New Haven County, CT (**Fig 4**). We observe that B.1.1.7 abundances predicted from wastewater are underestimated compared to the clinical abundance data. Based on our benchmarking experiments on whole genome data (**Fig 2**, left) we expect frequencies to be underestimated by 5-40% (relative to the actual frequency), with stronger underestimation for lower frequencies. This is consistent with what we see in **Figure 4**: the increase (and subsequent decrease) of B.1.1.7 abundance in wastewater is stronger than in clinical data because of this bias, while wastewater abundance predictions are very close to clinical predictions at frequencies of 60-70%. For B.1.526, both clinical and wastewater abundance is relatively stable over time. In theory, predictions will be closer to clinical abundances when sequencing only the Spike gene instead of the whole SARS-CoV-2 genome (**Fig 2**). In practice, however, using only reads aligning to the Spike gene captures too little information due to incomplete genome coverage and amplification bias.

Kallisto offers a bootstrapping feature, through which the sequencing data is resampled at least 100 times and variant abundances are predicted for each of these resampled datasets. The resulting predictions can subsequently be analyzed to obtain confidence intervals for the predicted abundance on the original dataset. For the New Haven sludge samples discussed here we obtained narrow confidence intervals (upper and lower errors <1% abundance), suggesting that predictions are very consistent (**Fig S6**). However, this type of analysis captures only computational noise, and not technical noise (e.g. sampling bias). The fact that our predictions are more variable than the clinical data while confidence intervals from bootstrapping are narrow suggests that wastewater sequencing data is highly stochastic and not always representative of the infections in the population.

### Wastewater abundances for different variants across the US match expected patterns

The quality of datasets obtained through wastewater sequencing varies widely. RNA levels and Ct values fluctuate with the number of infections in a sewershed, which impact overall genome coverage and prediction accuracy. Other factors such as sampling approach, PCR inhibition, and amplification bias add to this variability. To validate our approach for general use, we applied our pipeline to predict variant abundance on a diverse collection of composite influent wastewater samples obtained from 25 treatment plants across the United States between late December 2020 and late January 2021 (**Fig 5**). These samples had slightly higher average Ct values than the sludge samples analyzed above (33.1 vs 32.3), but reduced genome coverage compared to sludge samples. Nevertheless, the same quality filtering parameters apply: Ct < 31 or Ct < 34 with at least 0.5M reads aligned to select for samples with high coverage (**Fig S7**).

**Figure 5.**
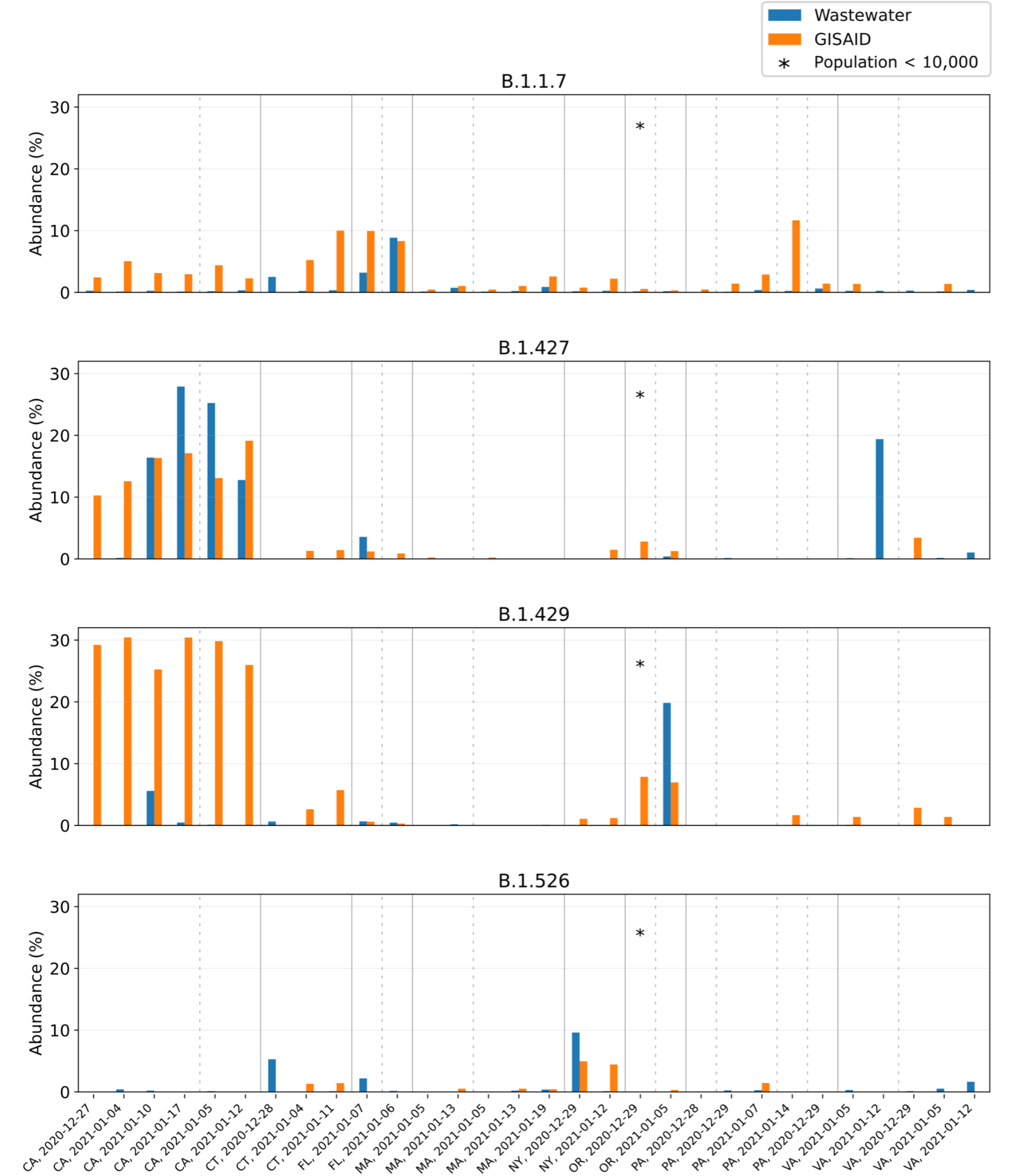
Wastewater versus GISAID abundance estimates for B.1.1.7, B.1.427, B.1.429 and B.1.526 at 16 locations across 8 states of the US. Samples were collected between late December 2020 and late January 2021; sampling date and location are indicated on the horizontal axis. Samples are sorted by location, with different locations separated by a dotted line and different states separated by a solid line.

After this filtering step, we predicted variant abundance for the 30 remaining datasets (corresponding to 16 different locations across 8 states). **Figure 5** shows the predictions for lineages B.1.1.7, B.1.427, B.1.429, and B.1.526, along with the clinical lineage frequencies in the corresponding state (calculated from GISAID in the 7-day window centered at the wastewater sampling date). Although data uploaded to GISAID has its own biases and individual towns do not necessarily reflect state-wide variant abundances, this is the only statistic we can compare against across all states. We observe that, while individual samples are unreliable, the predicted variant abundances match expected patterns across the US from the times of sampling: B.1.1.7 was predicted most abundantly in Florida; B.1.427 and B.1.429 were primarily found in California; and B.1.526 was predicted most abundantly in New York and Connecticut. Other variants (B.1.351, P.1) were not observed in GISAID for these states at the time of sampling and our predictions for these variants agree: B.1.351 was predicted to be present at very low frequency in 4 samples and absent in all other samples; P.1 was predicted present in a single dataset at 1% abundance and absent in all others (**Fig S8**). Although these predictions may be false positives, at the time P1 was thought to be likely at such low prevalence that these cases were not picked up by the sequencing efforts in place.

## Discussion

Our results show that methods for RNA transcript quantification can be applied to wastewater sequencing data to obtain consistent and relevant variant abundance estimates. This technique can be readily applied to a wide range of data types, from Spike-only amplicon sequencing to whole genome sequencing; it is not appreciably more difficult than setting up a “reference set” of potential variants and running existing tools on whatever the sequencing data happens to be. While this reference set approach allows easy updating as new variants appear, it also means that this approach cannot be used to detect new variants, but only to near-optimally impute the mixture of known variants most likely responsible for the observed data. Further, the approach may be readily extended to the detection and quantification of other pathogen lineages present in wastewater.

Detecting variants in this fashion appears to be near-optimal on simulated data, with a detection limit under 1%, though when handling real data which is subject to multiple factors that can alter the quality of the RNA and the resulting signal, this may be closer to 10%. Wastewater abundances generally follow the abundance trends seen in clinical data, though with sufficient noise that individual timepoints should not be considered reliable abundance estimates. Predicting variant abundance from wastewater sequencing is challenging when viral titers are low, as a result of low prevalence of SARS-CoV-2, but in high-prevalence regions this approach can be extremely effective.

When comparing wastewater abundances to population-level clinical frequencies of variants, there are three distinct potential sources of errors, all of which are conflated in the accuracy of the final estimate: (***1***) how well clinical frequencies match the rates in the population as a whole, (***2***) how representative the RNA in a given wastewater is of the infections in the population as a whole, and (***3***) how well the predicted variant abundances from sequencing accurately represent what is in the wastewater sample itself. Clinical frequencies and the GISAID data used for the national data above have strong biases and so are not themselves ground truth. Individual wastewater samples can be unreliable: the catchment and composition of an individual wastewater sample can include hospital or industrial inputs and is not necessarily representative of the population, and low infection levels, inhibitory compounds, and degradation of RNA can result in higher Ct values and associated genome coverage. Further, the potential that different variants have different viral shedding in waste is not taken into account nor are differences in shedding rates for vaccine breakthrough cases. The data here is consistent with high-coverage sequencing being well representative of the sample and the computation faithfully reconstructing it, therefore we believe the primary source of observed noise is the underlying noisiness correspondence between a given wastewater sample, the population as a whole, and the clinical cases.

These results do offer lessons for future development of wastewater sequencing methods. With perfect coverage (simulated data), Spike-only sequencing gives better predictions than whole genome sequencing. More broadly, as the variant-discriminating mutations are restricted to a subset of the amplicons in tiled sequencing, a strategy focusing sequencing depth proportionally to the potentially informativity of the tiles would likely yield the more accurate predictions per sequence read. In practice, whole genome sequencing is often necessary to obtain enough information versus restricting to Spike. These differences in sequencing protocols, as well as differences in population size and sampling techniques, make it challenging to compare data between locations. Another important lesson is the improved coverage with higher virus RNA concentrations (lower PCR Ct values). While Ct is largely controlled by the infection rate in a community and excretion into wastewater, sample concentration and PCR inhibition removal approaches can be deployed to lower the Ct value and improve coverage in most samples.

In applying RNA-seq tools to variant prediction, reference set composition is central. We were able to reduce variant-specific biases substantially by including multiple reference sequences per lineage, thus capturing within-lineage variation to identify variants with highly similar genomes. Additional experiments (data not shown) comparing a US-specific reference set to a global reference set showed that, unsurprisingly, the US-specific reference set gives more accurate predictions for benchmarks with sequences from US origin. This suggests that using a state- or county-specific reference set construction could further improve results and that identification of variants is likely being aided by the presence of hitchhiker mutations which are locally over-represented or non-defining.

In conclusion, we present a computational approach for estimating the percent abundances of SARS-CoV-2 variants in wastewater. Temporal patterns in wastewater of variant abundances in a mid-size municipality in Connecticut matched those defined by compiled clinical sequence data, and sequencing metrics were interrogated to define the Ct value and other confidence thresholds that ensure optimal performance. In settings across the world where strong clinical variant sequencing programs do not exist, wastewater sequencing can be an effective tool for low cost, efficient monitoring of variant abundance. As this is unlikely to be the last viral pandemic, nor the last with variants of concern, extending these approaches to other viruses and other sample types may allow broader monitoring of real-time pandemic evolution.

## Methods

### Constructing a reference set

We selected representative genomes per lineage from the GISAID database^4^, downloaded on 9 March 2021. As our samples are from wastewater collection systems across the US, we considered only reference sequences of US origin. After removing low-quality sequences (defined as having less than 29,500 non-ambiguous nucleotides) we randomly selected 1000 sequences per lineage for further analysis. We used minimap2 and paftools to align each of these sequences to the reference genome (MN908947.3) and subsequently identify variation with respect to this reference^22^. We then used VCFtools to compute allele frequencies within each lineage. Based on these allele frequencies, we selected sequences per lineage such that all mutations with an allele frequency of at least 50% were captured at least once. This resulted in a final reference set of 1,488 complete SARS-CoV-2 genome sequences.

### Designing benchmarking experiments

To evaluate the accuracy of our variant abundance predictions, we created benchmarks consisting of a selection of non-variant genomes (background) and one variant. In order to build benchmarks that reflect our real data as closely as possible, we selected the background genomes by taking all 11 sequences in GISAID collected in Connecticut at 2021-02-11 (the most recent collection date). For Spike-only benchmarks, we trimmed these sequences to keep only the Spike gene and simulated paired-end (2×150 bp) Illumina sequencing reads at equal abundance using ART^23^. In addition, we randomly selected variant sequences from GISAID and simulated sequencing reads for the Spike gene of each variant (B.1.1.7, B.1.351, P.1, B.1.427, B.1.429) at varying frequencies (0.05, 0.06 …, 0.1, 0.2, …, 1, 2, …, 10, 20, …, 100%) to create 33 data sets per variant, hence 165 data sets in total. We performed these simulations at a total coverage of 100x and 1000x for whole genome benchmarks, and at 100x, 1000x, and 10,000x for Spike-only benchmarks.

### Wastewater collection and sequencing from New Haven, CT

Primary sewage sludge samples were collected from the New Haven, CT, USA Wastewater Treatment Plant. The plan serves 200,000 residents in the towns of New Haven, Hamden, East Haven and Woodbridge, CT. Primary sludge samples were collected from the effluent pump of the plant’s gravity thickener. Samples were collected every other day starting January 3, 2021 and ending April 27, 2021. RNA was extracted from 500 µL sample using a Zymo *Quick*-RNA Fecal/Soil Microbe Microprep Kit modified by the addition of 100 µL of phenol-chloroform to the bead beating step, and eluted in 50 µL of nuclease-free water. The SARS-CoV-2 whole-genome sequencing library was prepared from extracted RNA using the Illumina COVIDSeq Test (RUO), modified to use NEBNext ARTIC V3 SARS-Cov-2 Primer Mixes 1 and 2 instead of the included primer mixes, for cDNA synthesis, amplicon generation, tagmentation, and cleaning. The pooled and cleaned library was sequenced on an Illumina NovaSeq at the Yale Center for Genomic Analysis; each sample was given at least 1 million reads. Negative controls were included at the cDNA synthesis and amplicon generation steps.

### Wastewater collection and sequencing from across the US

Composite influent samples were collected by participating wastewater treatment facilities using equipment that these facilities already had in-house. Composite samples were aliquoted into three 50-mL conical tubes and shipped within 24 hours of collection overnight with ice packs to the Biobot Analytics laboratory (Cambridge, MA). Received samples were immediately pasteurized at 60°C for 1h.

One of the three tubes was then filtered to remove large particulate matter using a 0.2uM vacuum-driven filter (EMD-Millipore SCGP00525 or Corning 430320, depending on sample turbidity). We then used Amicon Ultra-15 centrifugal ultrafiltration units (Millipore UFC903096) to concentrate 15mL of wastewater approximately 100x. We lysed viral particles in the concentrate by adding AVL Buffer containing carrier RNA (Qiagen 19073) to the Amicon unit before transfer and >10 minute incubation in a 96-well 2mL block. To adjust binding conditions, 100% ethanol was added to the lysate, and samples were applied to RNeasy Mini columns or RNeasy 96 cassettes (Qiagen 74106 or 74181). For a subset of samples (all from locations within Massachusetts) we processed 45mL of wastewater by loading the same Amicon Ultra-15 unit three times.

The RNA samples resulting from the extraction process described above were used as the template for reverse-transcription (RT) reactions performed with LunaScript RT SuperMix enzyme mix (NEB) to generate cDNA. Reaction conditions were as follows: primer annealing at 25°C for 2 min, cDNA synthesis at 55°C for 10 min and heat inactivation at 95°C for 1 min. Multiplexed polymerase chain reaction (PCR) amplification of cDNA was performed with Q5 Hot Start High-Fidelity 2X Master Mix (NEB) and ARTIC v3 primers (0.015 µM each, final) in two non-overlapping pools with the following cycling conditions: heat activation at 98°C for 30 sec, followed by 35 cycles of 15 sec denaturation at 98°C, 5 min annealing/elongation at 65°C.

The non-overlapping amplicon pools were combined and sequencing libraries for Illumina platform were prepared using tagmentation with bead-linked transposomes (Illumina) and a modified amplification protocol with KAPA HiFi HotStart ReadyMix (Roche) and combinatorial dual-indexed adapter sequences. Libraries were sequenced with NextSeq550 (Illumina).

### Wastewater data preprocessing

Before processing with kallisto, we first removed adapter sequences from the reads using Trimmomatic^24^, aligned the trimmed reads to a reference genome (GenBank MN908947.3) with BWA-MEM v0.7.17^25^, and subsequently identified primer sequences using iVar v.1.3.1^16^ and removed these with jvarkit (http://lindenb.github.io/jvarkit/Biostar84452).

### Clinical sequencing and data processing from New Haven, CT

#### Ethics statement

The Institutional Review Board from the Yale University Human Research Protection Program determined that the RT-qPCR testing and sequencing of de-identified remnant COVID-19 clinical samples obtained from clinical partners conducted in this study is not research involving human subjects (IRB Protocol ID: 2000028599).

#### Sequencing and consensus generation

Residual routine testing samples from confirmed SARS-CoV-2 positive individuals were provided by Yale New Haven Hospital, Yale Pathology Laboratory, “Yale Campus Study”, Connecticut Department of Public Health, and Murphy Medical Associates. Sample types included nasal swabs in viral transport media, raw saliva, and extracted RNA. Samples not arriving as RNA were processed using the MagMAX viral/pathogen nucleic acid isolation kit; RNA was extracted from 300 µL of sample and eluted in 75 µL elution buffer. All products were tested using a locally developed assay for variants to determine viral RNA concentration^26^. Samples with sufficient RNA for sequencing (defined as a viral target cycle threshold value <35) were prepared using the Illumina COVIDSeq Test RUO for cDNA synthesis, amplicon generation, tagmentation, and cleaning. Pooled and cleaned libraries were sequenced using a 2×100 or 2×150 approach on an Illumina NovaSeq at the Yale Center for Genomic Analysis; each sample was given at least 1 million reads. Negative controls were included at RNA extraction, cDNA synthesis, and amplicon generation steps.

Reads were aligned to a reference genome (GenBank MN908937.3) using BWA-MEM v.0.7.15^25^. Adaptor trimming, primer sequence masking, and simple majority base calling were conducted using iVar v1.2.1^16^ and SAMtools^27^. Lineages were assigned using pangolin v.2.4.2^28^.

### Software availability

All code used for the analysis presented in this manuscript is publicly available at https://github.com/baymlab/wastewater_analysis.

## Data Availability

The raw SARS-CoV-2 sequencing data from New Haven wastewater (.fastq files) are available on NCBI SRA under Bioproject PRJNA741211. The clinical sequencing data can be accessed via covidtrackerct.com. The raw SARS-CoV-2 sequencing data from across the U.S. (.fastq files) are available on NCBI SRA under Bioproject PRJNA759260. The simulated wastewater sequencing data (.fastq files) for benchmarking are available on Zenodo (DOI: 10.5281/zenodo.5307070).

https://www.ncbi.nlm.nih.gov/bioproject/PRJNA741211

https://www.ncbi.nlm.nih.gov/bioproject/PRJNA759260

https://zenodo.org/record/5307070#.YS6eZNNKjUI

## Yale SARS-CoV-2 Genomic Surveillance Initiative authors

Ahmad Altajar, Anderson F. Brito, Anne E. Watkins, Anthony Muyombwe, Caleb Neal, Chen Liu, Christopher Castaldi, Claire Pearson, David R. Peaper, Eva Laszlo, Irina R. Tikhonova, Jafar Razeq, Jessica E. Rothman, Jianhui Wang, Kaya Bilguvar, Linda Niccolai, Madeline S. Wilson, Margaret L. Anderson, Marie L. Landry, Mark D. Adams, Pei Hui, Randy Downing, Rebecca Earnest, Shrikant Mane, Steven Murphy

## Acknowledgements

This work was supported in part by the Pew Charitable Trusts, the David and Lucile Packard Foundation, NIH NIGMS award R35GM133700, and the Alfred P. Sloan Foundation (J.A.B. and M.B); CTSA Grant Number TL1 TR001864 (M.E.P. and T.A.); Fast Grant from Emergent Ventures at the Mercatus Center at George Mason University (N.D.G.); CDC Contract #75D30120C09570 (N.D.G.); Yale CoReCT pilot award (J.P. and N.D.G.); and NIH NIGMS award U54GM088558 (W.P.H.). James McGann and Jim Griffin were involved in developing the sequencing methodology at Gingko Bioworks.

## Supplementary figures

**Figure S1:**
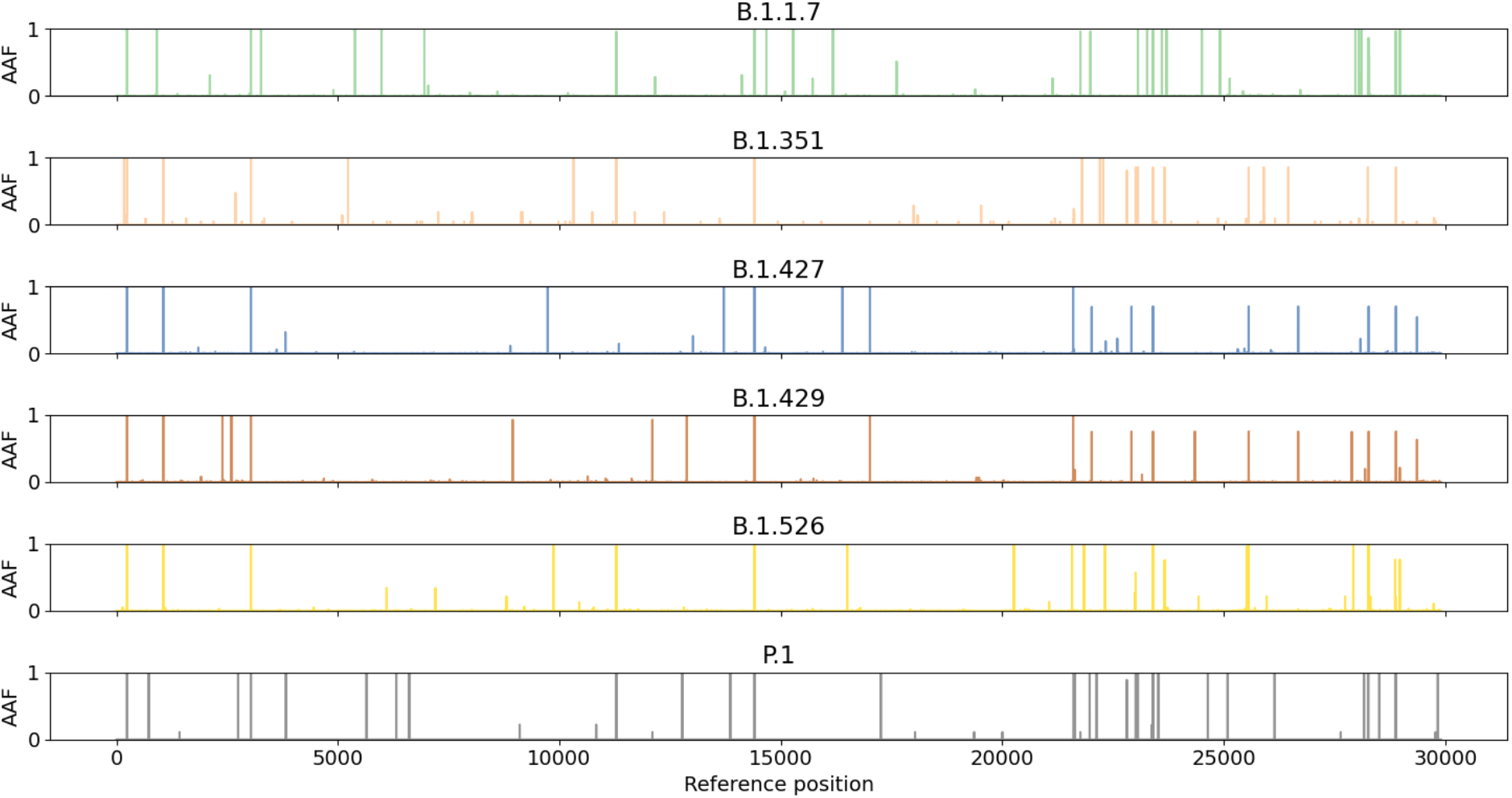
Within-lineage diversity observed in SARS-CoV-2 genomes on GISAID (downloaded 9 March 2021). The horizontal axis shows the position (in base pairs) on the reference genome (accession MN908947.3). The y-axis shows the alternative allele frequency (AAF), i.e. the fraction of genomes with a different nucleotide at a given position than the reference genome. This plot was computed by randomly selecting 1000 genomes of US origin per lineage.

**Figure S2.**
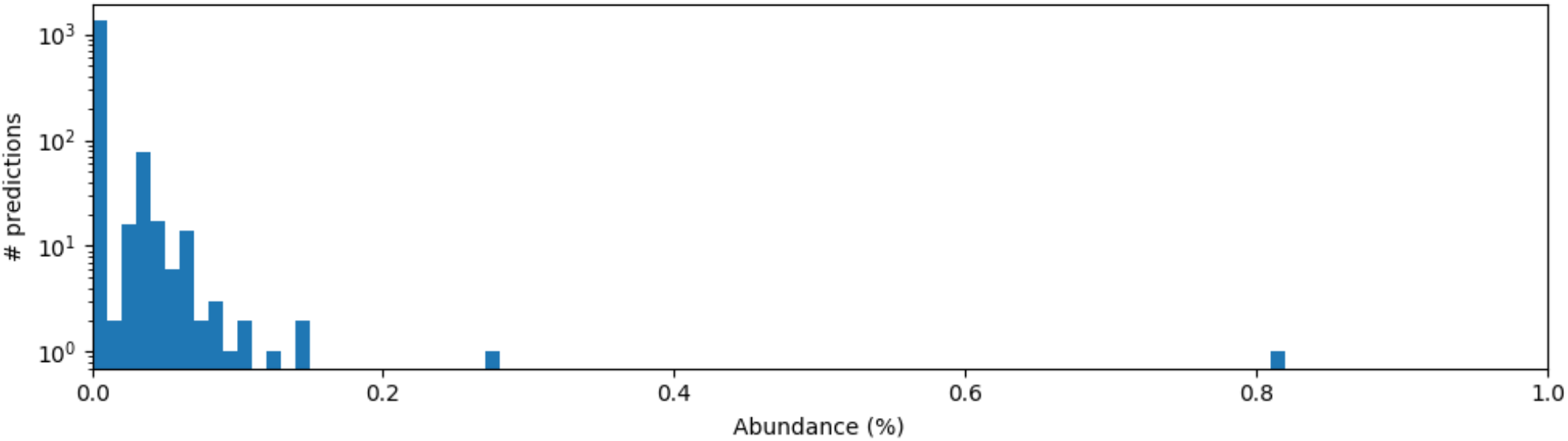
Histogram of raw abundances predicted by kallisto on a simulated dataset consisting of 100% B.1.1.7. The majority of false positives (background noise) can be filtered out by applying a minimal abundance threshold of 0.1%. True predictions occur at higher abundances (beyond the x-axis limit of 1.0%).

**Figure S3:**
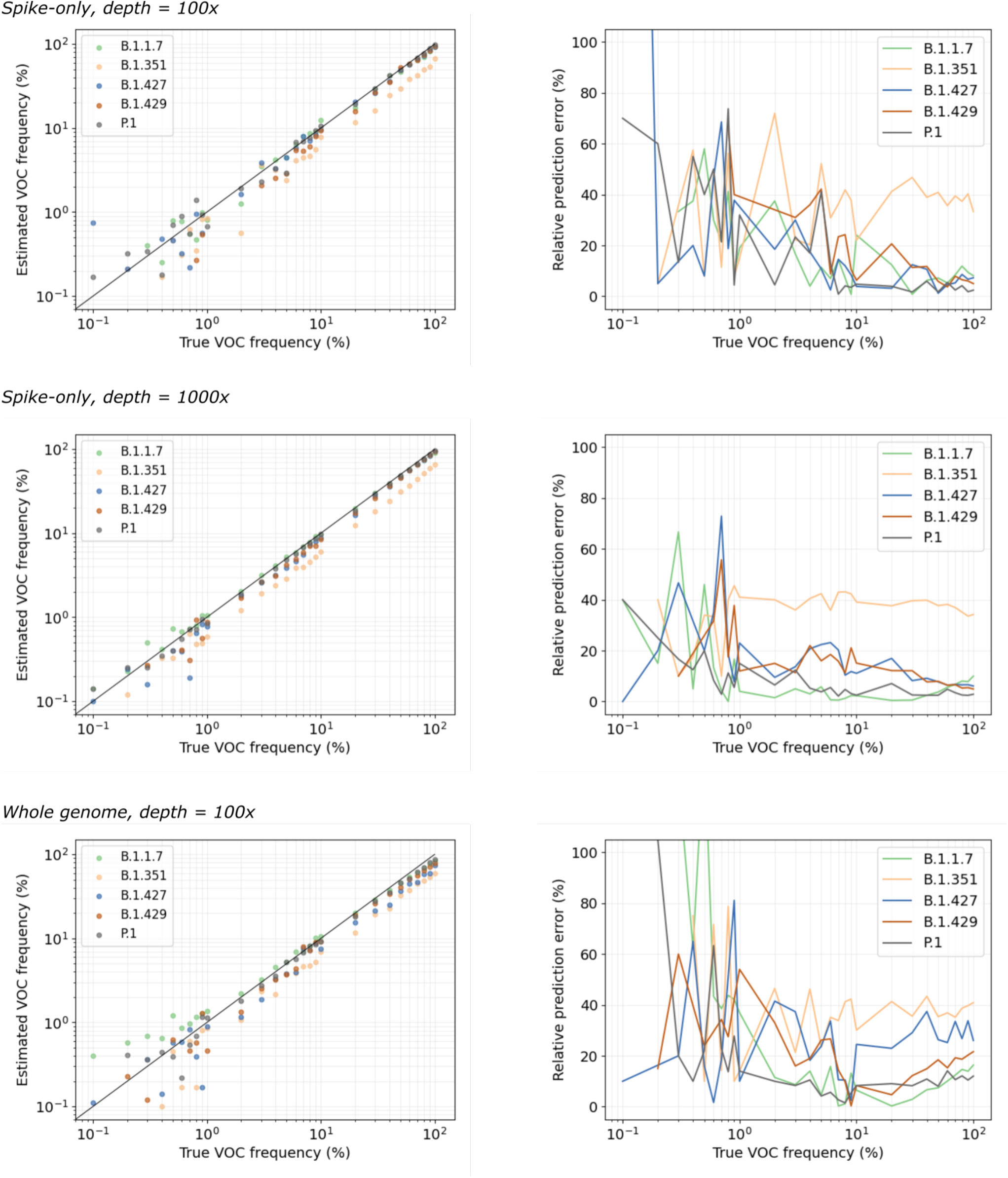
Additional benchmarking results

**Figure S4:**
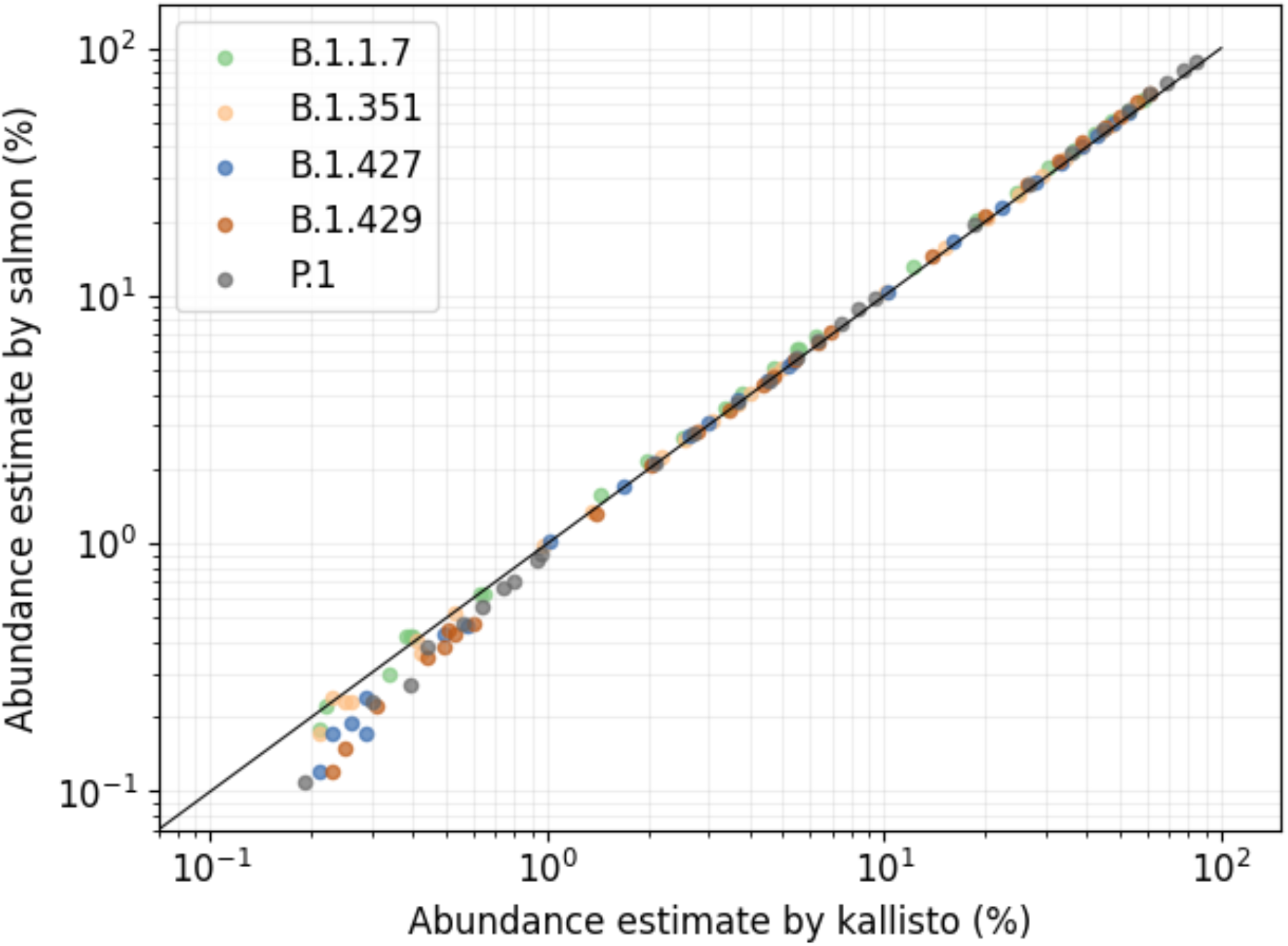
salmon versus kallisto abundance estimates per variant.

**Figure S5:**
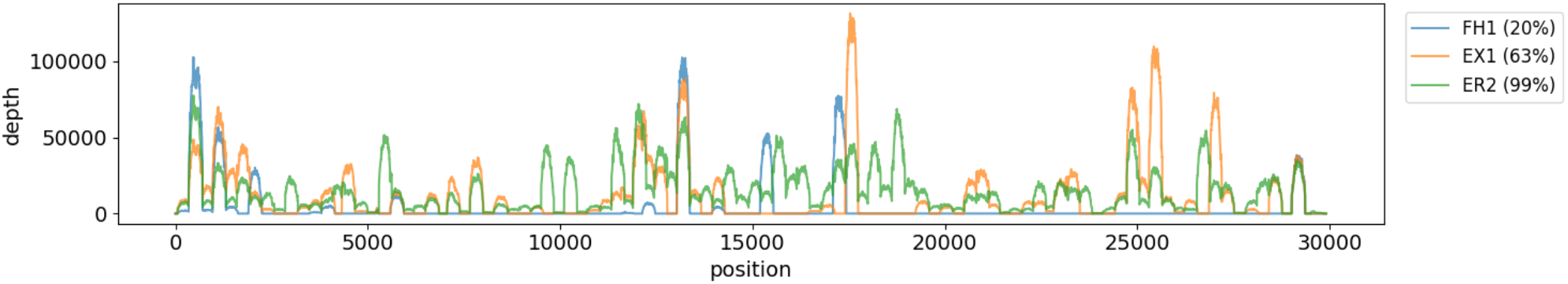
Sequencing depth from wastewater samples is highly uneven between amplicons. This figure shows sequencing depth along the genome for three samples collected in New Haven, CT. The first sample (FH1) has low genome coverage (20%), with very few amplicons reaching high sequencing depth. The second sample (EX1) has moderate genome coverage (63%), with roughly half of the amplicons reaching high sequencing depth. The third sample (ER2) has high genome coverage (99%), with nearly all amplicons reaching high sequencing depth.

**Figure S6:**
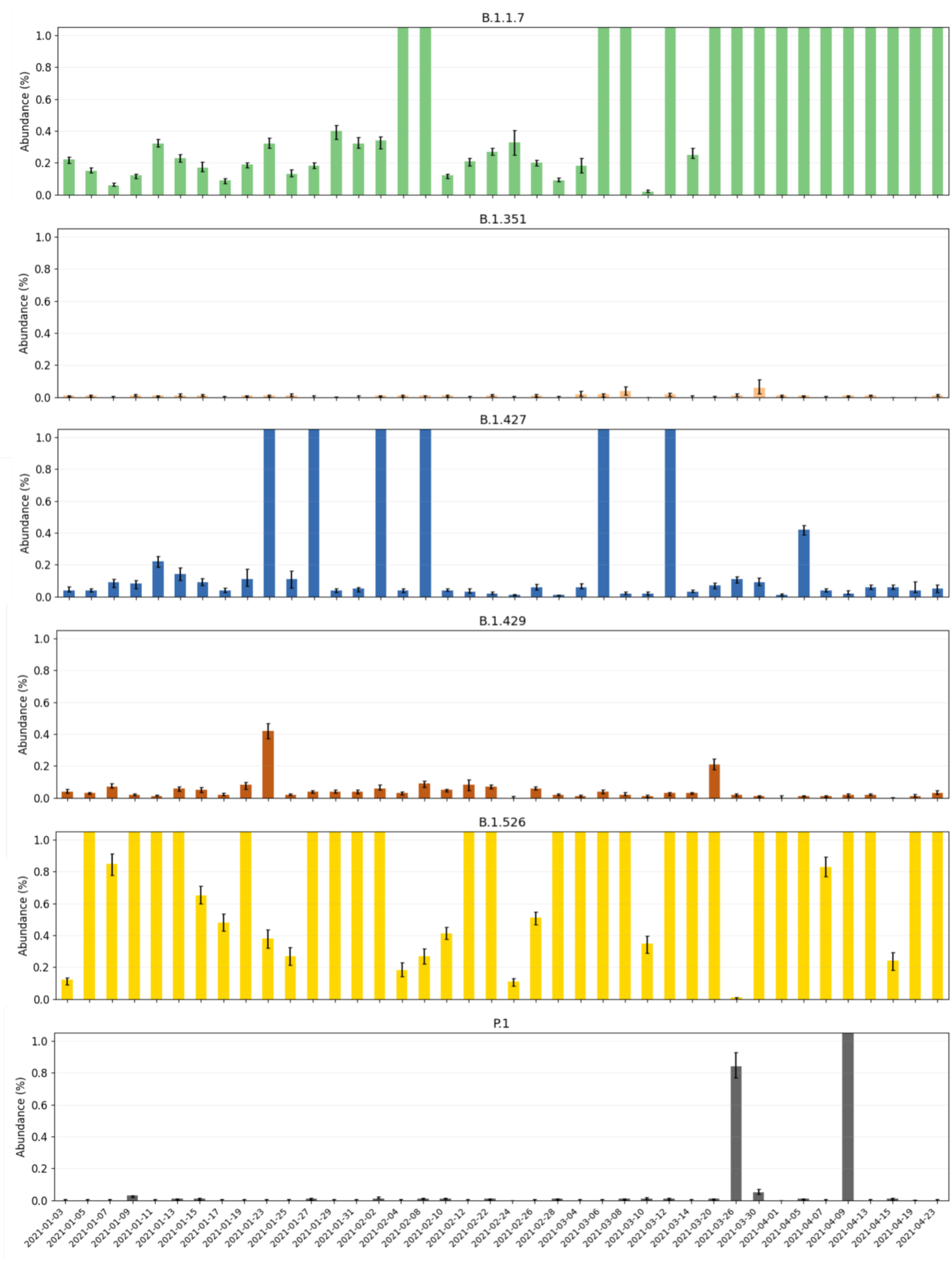
Raw predictions per variant with confidence intervals based on bootstrap analysis for New Haven samples. Note that in all subplots the y-axis is capped at 1% for improved readability.

**Figure S7:**
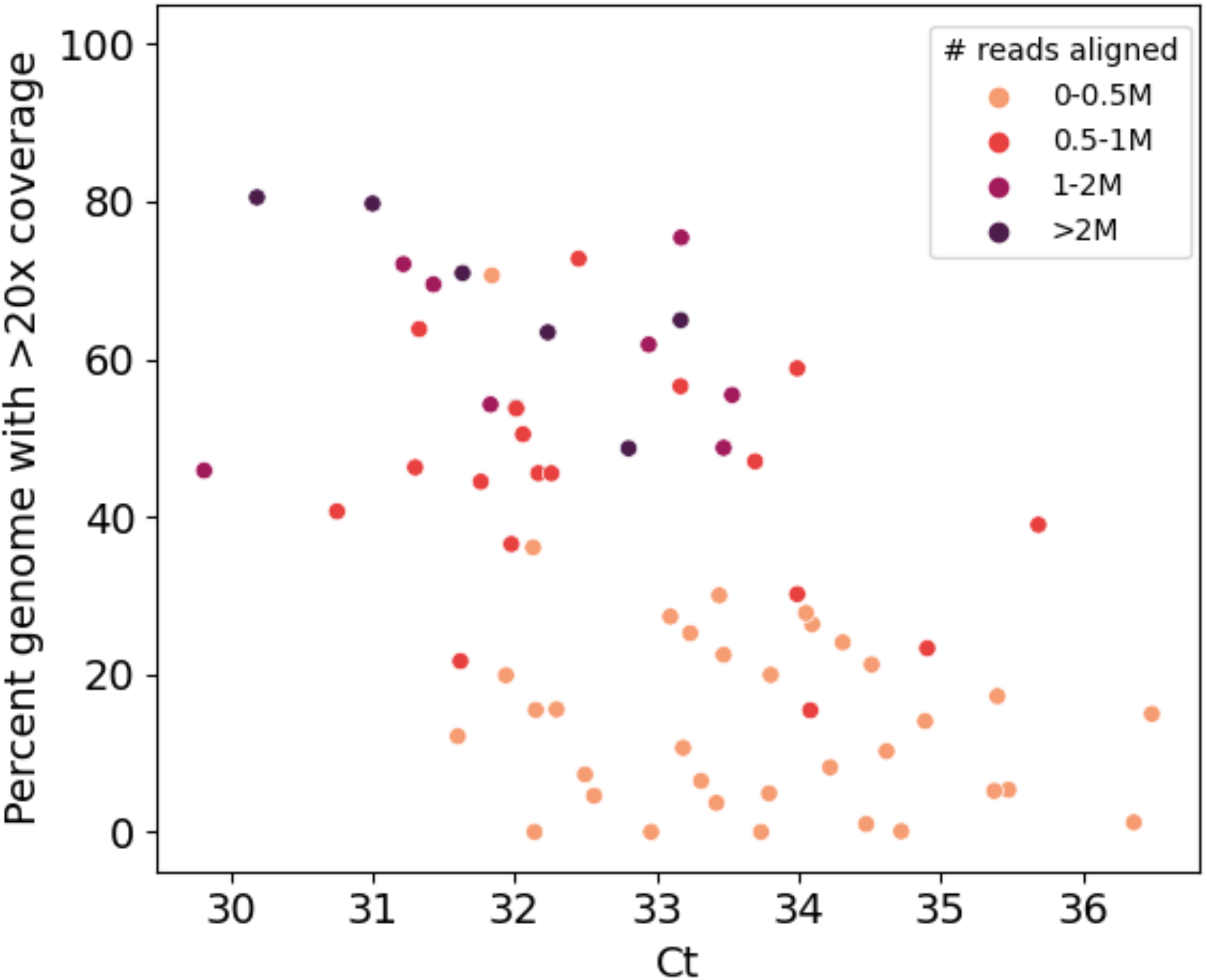
Percent genome coverage versus Ct values for samples across the US

**Figure S8:**
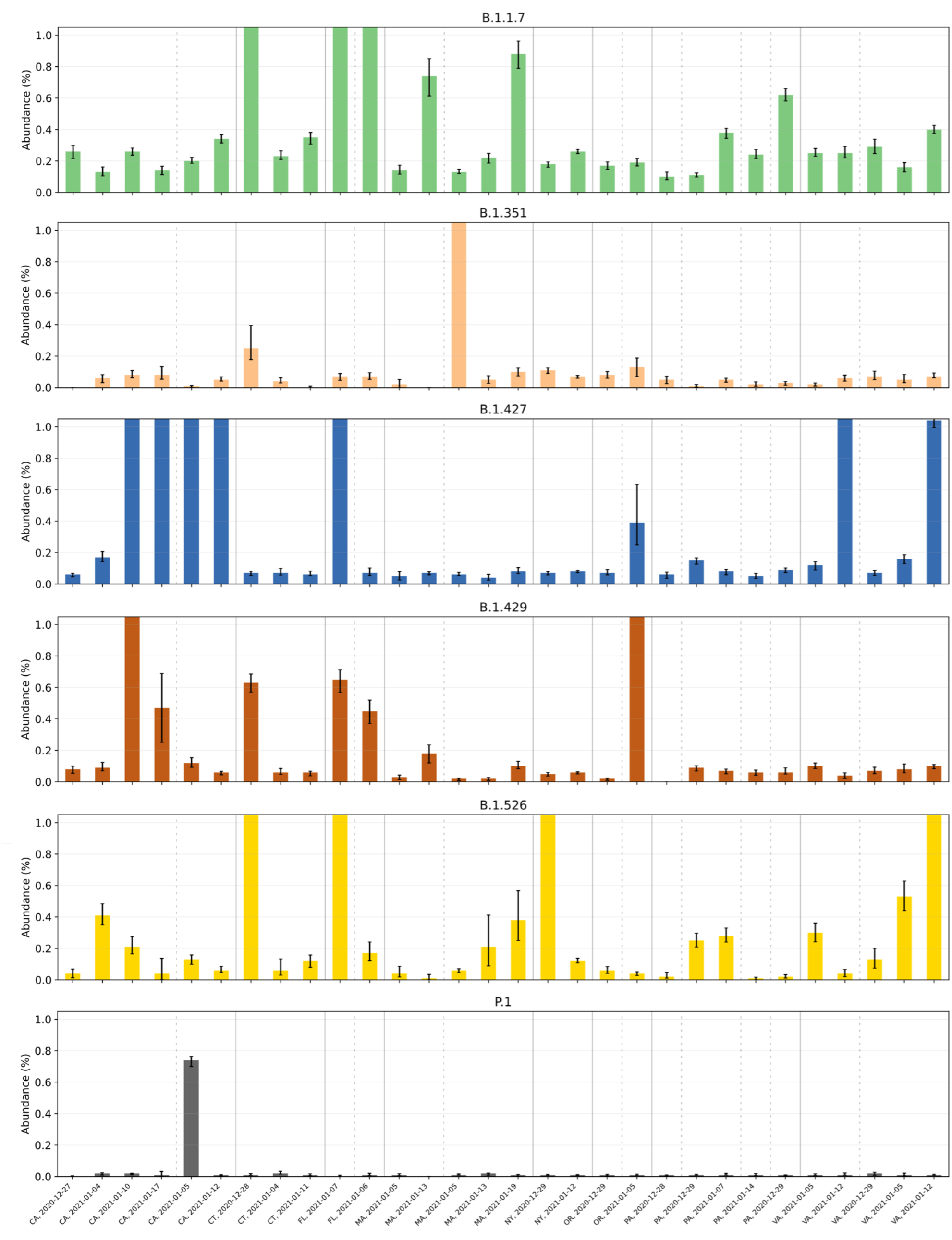
Raw predictions per variant with confidence intervals based on bootstrap analysis for samples across the US. Note that in all subplots the y-axis is capped at 1% for improved readability.

